# Efficacy and Safety of Tradipitant in Diabetic and Idiopathic Gastroparesis: A Randomized, Placebo-Controlled Study

**DOI:** 10.1101/2020.04.10.20057976

**Authors:** Jesse L. Carlin, V. Rose Lieberman, Arya Dahal, Madison S. Keefe, Changfu Xiao, Gunther Birznieks, Thomas L. Abell, Anthony Lembo, Henry P. Parkman, Mihael H. Polymeropoulos

## Abstract

**Background and Aims:** There is a high unmet need for the treatment of gastroparesis and studies of NK1-R antagonists suggest potential benefit in reducing the symptoms of nausea and vomiting. We hypothesized that tradipitant, an NK1-R antagonist, would be effective in treating patients with idiopathic or diabetic gastroparesis.

**Methods:** In a randomized, double-blind, placebo-controlled study across 47 U.S. sites, 152 gastroparesis patients were randomized to receive oral 85mg BID tradipitant (n=77) or placebo (n=75) daily for four weeks. Symptoms were assessed using a daily symptom dairy, Gastroparesis Cardinal Symptom Index (GCSI), and other patient reported questionnaires.

**Results:** Patients receiving tradipitant had a significant decrease in nausea score at Week 4 compared to placebo (−1.2 improvement vs −0.7, respectively, p=0.0099), and a significant increase in nausea-free days (28.8% increase on tradipitant vs 15.0% on placebo p=0.0160). Patients with both nausea and vomiting at baseline (n=101) showed an even greater decrease in nausea score (−1.4 improvement on tradipitant vs −0.4 on placebo p<0.0001) and an increase in nausea free days (32.3% improvement on tradipitant vs 7.6% on placebo p=0.0003). 32.9% of patients treated with tradipitant were nausea responders (average nausea score ≤ 1 at week 4) compared to 11.8% of patients on placebo (p=0.0013). 46.6% of patients treated with tradipitant had a greater than 1-point improvement in GCSI score compared to 23.5% of patients on placebo (p=0.0053).

**Conclusions:** Tradipitant treatment resulted in statistically and clinically meaningful improvements in nausea and overall gastroparesis symptoms. These robust efficacy results suggest tradipitant has the potential to become a useful pharmacological treatment for gastroparesis.

## Introduction

Gastroparesis is a serious chronic medical condition characterized by delayed gastric emptying. Gastroparesis is frequently associated with significant impairment of social and occupational functioning. The symptoms of gastroparesis include nausea, vomiting, bloating, fullness after meals, and abdominal discomfort and pain. Nausea is the most common symptom of gastroparesis and is reported in more than 90% of individuals with gastroparesis. Vomiting and bloating, other major symptoms of gastroparesis, are reported in 68-84% and in 75% of patients respectively^1^.

The incidence of gastroparesis ranges from 6.3 to 17.2 per 100,000 person-years^2^. The prevalence of gastroparesis has been reported to be 24.2 per 100,000; however, because gastroparesis is underdiagnosed, the true prevalence has been estimated to be 50.5 per 100,000 and therefore may affect over 5 million individuals in the United States, which is approximately 1.5% of the population^3^.

The pathophysiology of gastroparesis is complex and probably involves neuromuscular dysfunction and sensory neuropathy resulting in delayed gastric emptying, nausea and pain. It is recognized that treatment should target symptoms of gastroparesis and not only gastrointestinal motility due to the weak relationship between upper GI symptoms and the rate of gastric emptying^4,5,6^. There is a high, unmet medical need for gastroparesis therapies for chronic use.

The only U.S. Food and Drug Administration approved treatment for gastroparesis is metoclopramide, which carries a black box warning and limitations of use of no more than 3 months due to the serious neurological side effect of tardive dyskinesia. Clinical guidelines recommend, in addition to metoclopramide, the off label use of erythromycin, domperidone (not approved in the U.S.), botulinum toxin injections, gastric stimulators, and a variety of surgical procedures in an effort to relieve some of the symptoms of the disease^7,8^. The current treatment landscape for gastroparesis is in critical need for solutions and currently includes drugs with high rates of side effects, limited use restrictions, or invasive procedures with limited evidence of their efficacy^7^.

NK-1 receptor (NK-1R) antagonists are approved to treat nausea and vomiting in chemotherapy and the mechanism has been investigated in gastroparesis where it showed positive effects on nausea^9^. NK-1R antagonists may have a dual and potentially therapeutic effect in gastroparesis by affecting gastric motility through a local action as well as a direct effect in the brain regions responsible for nausea and vomiting. Neuronal signals from the gut or chemical signals from the blood are sensed in the brain to trigger nausea and vomiting. Substance P (SP) acts on the NK-1R and is believed to exert a key role within the central emetic circuitry along with serotonin ^10^. SP is also located in vagal afferents and in both the nerves and the muscular layer of the gastrointestinal tract. SP binds the NK-1R at the gastric neuromuscular junction where there is a functional interplay between the acetylcholine and NK-1R systems to stimulate smooth muscle contractions^11^.

Tradipitant (VLY-686) is a potent selective inhibitor of NK-1R and is hypothesized to treat gastroparesis by acting centrally in the nausea-vomiting centers of the brain and peripherally in the smooth muscle of the intestines. Here we report results of a 4-week, multi-center, double-blind, placebo-controlled, randomized clinical trial comparing 85mg tradipitant BID with placebo in idiopathic and diabetic gastroparesis.

## Methods

The safety and efficacy of tradipitant to treat gastroparesis symptoms was assessed in a 4-week multi-center, double-blind, placebo controlled, randomized trial of patients with idiopathic or diabetic gastroparesis and moderate to severe nausea (VP-VLY-686-2301). Patients were randomized to treatment with either tradipitant capsules (85mg twice daily) or placebo capsules (twice daily) and stratified by disease etiology (diabetic or idiopathic). The primary outcome from the intent-to-treat (ITT) analysis was change from baseline to Week 4 (Day 22 to 28) in average nausea severity as measured by the Gastroparesis Core Symptom Daily Diary (GCSDD).

### Trial Design

This phase II trial took place in 47 sites across the United States from November 2016 until December 2018 (NCT02970968). Authors had access to the study data and had reviewed and approved the final manuscript.

All protocols, source documents, and patient-facing material was reviewed and approved by the independent Institutional Review Board, Advarra (Maryland, USA). All subjects were informed of study procedures and risks by a qualified study team member before any study procedures were performed. All subjects signed an informed consent form and were given a copy to keep.

Eligible patients aged 18-70 years with a diagnosis of gastroparesis as demonstrated by delayed stomach emptying and symptoms of gastroparesis (nausea, early satiety, fullness, etc.) entered a 4-week screening period. At the conclusion of the screening period, patients meeting all criteria were randomly assigned (1:1) treatment to tradipitant or placebo for 4 weeks. Patients were seen at Day −28, −14, 0, 14 and 28.

### Inclusion and Exclusion Criteria

Patients were required to have moderate to severe nausea during the 4-week screening period defined as an average nausea score for the worst 50% of their screening days of ≥3 on a 5 point scale. Patients were excluded if they used narcotics more than 2 times per week, used domperidone or other experimental medications in the last 60 days, had a gastric stimulator implanted in the last year, or setting changed in the last 3 months, had a gastric surgery (i.e. gastric bypass, gastrectomy, pyloroplasty), or had elevated alanine aminotransferase (ALT) or aspartate aminotransferase (AST) at 1.5 times the upper limit of normal. Patients on stable doses of other gastrointestinal or anti-emetic drugs, including metoclopramide, erythromycin, ondansetron, prochlorperazine, and promethazine were not excluded and patients were allowed to remain on these medications. Ondansetron, prochlorperazine and promethazine were allowed as rescue medication for nausea throughout the entire study and dosing was recorded. Patients had to demonstrate evidence of delayed gastric emptying within the last 10 years. If patients did not have a gastric emptying study in the last 10 years, delayed gastric emptying was confirmed via a gastric emptying breath test (Cairn Diagnostics, Brentwood, TN).

### Assessments

Patients completed a daily patient reported symptom diary, Gastroparesis Core Symptom Daily Diary (GCSDD), for a 4-week (28 day) screening period and during the 4-week treatment period. In addition, the following questionnaires were administered at clinic visits: Patient Assessment of Gastrointestinal Disorders Symptom Severity Index (PAGI-SYM) which includes the Gastroparesis Cardinal Symptom Index (GCSI), Clinician Global Impression of Severity (CGI-S), Patient Global Impression of Change (PGI-C) and the Patient Assessment of Upper Gastrointestinal Disorders-Quality of Life (PAGI-QOL) questionnaire.

The PAGI-SYM is a patient reported outcome which asks patients to describe the severity of their symptoms over the last two weeks. The PAGI-SYM was developed to measure symptom severity for gastroparesis, functional dyspepsia, and gastroesophageal reflux disease^12^. The measure consists of 20 symptom severity items, which cover the following domains: nausea/vomiting, fullness/early satiety, bloating, upper abdominal pain, heartburn/regurgitation, and lower abdominal pain. The questionnaire uses a 0-5 Likert scale from 0 =none to 5 = very severe and includes the GCSI^12^. The GCSI total score was computed as the average of the following subscores: nausea/vomiting (3 items), fullness/early satiety (4 items) and bloating (2 items).

The PGI-C is a patient reported questionnaire with a 7 point rating scale where the subject rates their own improvement in overall symptoms relative to the baseline assessment. It is rated as: 1, very much improved; 2, much improved; 3, minimally improved; 4, no change; 5, minimally worse; 6, much worse; or 7, very much worse ^13^

The PAGI-QOL is a 30-item instrument assessing quality of life in patients with gastroparesis. The questionnaire covers five domains: Daily Activities, Clothing, Diet and Food Habits, Relationship, and Psychological Well-Being and Distress^14^. Effect on overall quality of life and well-being over the past two weeks is rated on a scale of 0 = “None of the time” to 5 “All of the time” for each item.

The GCSDD daily symptom diary asked patients to rate the worst occurrence of each cardinal symptom of gastroparesis in the past 24 hours on a 0 (no symptoms) to 5 (very severe) scale. This scale was based off of the ANMS GCSI-DD which has 5 questions rated on a 0-4 scale and has been previously validated in idiopathic and diabetic gastroparesis patients^15^. Consistent with the GCSI-DD available through the Mapi Research Trust (Lyon, France) and the 2-week GCSI site-based questionnaire, the GCSDD allowed patients to select the category “very mild” which is missing from the updated ANMS GCSI-DD.

As with ANMS GCSI-DD, the GCSDD included questions that address nausea ⁄ vomiting (3 items), postprandial fullness⁄ early satiety (4 items), and abdominal pain (2 items). We also added questions to the GCSDD about symptoms of bloating (2 items), hours of nausea, and the frequency of daily rescue medication use, which are not included in the original GCSI. Similar daily symptom scales based off the GCSI and the PAGI-SYM have been validated in gastroparesis patients.^16^ The full GCSDD utilized in the study can be found in Supplementary Material (S1).

Clinicians completed the CGI-S, which is a 7-point scale that the clinician rates the severity of the patient’s gastroparesis at the time of assessment and refers to the degree of illness at the time of the visit and during the two weeks prior to the visit^13^. The CGI-S is rated on the following seven-point scale: 1=normal, not at all ill; 2=borderline ill; 3=mildly ill; 4=moderately ill; 5=markedly ill; 6=severely ill; 7=among the most extremely ill patients.

### Statistical Analysis

All primary and secondary analyses were conducted on the intent-to-treat (ITT) population that included patients who were treated for at least 2 weeks (14 days). The primary and secondary outcomes were analyzed using a restricted maximum likelihood (REML)-based MMRM. The MMRM model included the fixed, categorical effects of treatment group, disease type, week, treatment group by week interaction, and pooled site as well as the fixed, continuous covariates of the baseline score and the baseline score by week interaction. A post-hoc Baseline Vomiting Group subpopulation consisted of 101/141 (72%) patients of the ITT Population. Additional post-hoc analyses were performed as responder analyses on GCSI, nausea, and nausea-free days as well as an anchor based analysis against PGI-C. Subject number was determined from sample size calculation with 86% power to detect a mean difference in nausea severity in a two-sided t-test with an alpha level of 0.05. All data processing, summarization, and analyses were performed using SAS^®^ version 9.3 or higher (SAS Institute Inc., Cary, NC). All authors reviewed study data and reviewed and approved the final manuscript.

## Results

### Patients

Between November 2016 and December 2018, 446 patients were screened and 152 patients were enrolled at 47 participating study sites. Of the enrolled patients, 77 were randomized to tradipitant and 75 to placebo. Twelve patients withdrew during the evaluation phase (initial 2 weeks of treatment) (tradipitant n=5, placebo n=7). Therefore, the ITT population included 141 patients (73 patients receiving tradipitant and 68 patients receiving placebo). The reasons for the withdrawals are shown in Figure 1. The Baseline Vomiting group included 101 (72%) patients (i.e., patients who reported at least one episode of vomiting during the screening period; tradipitant = 58; placebo = 43). 86 study patients were diagnosed with idiopathic gastroparesis (59.9%) and 55 patients were diagnosed with diabetic gastroparesis (40.1%, Table 1).

**Table 1.**
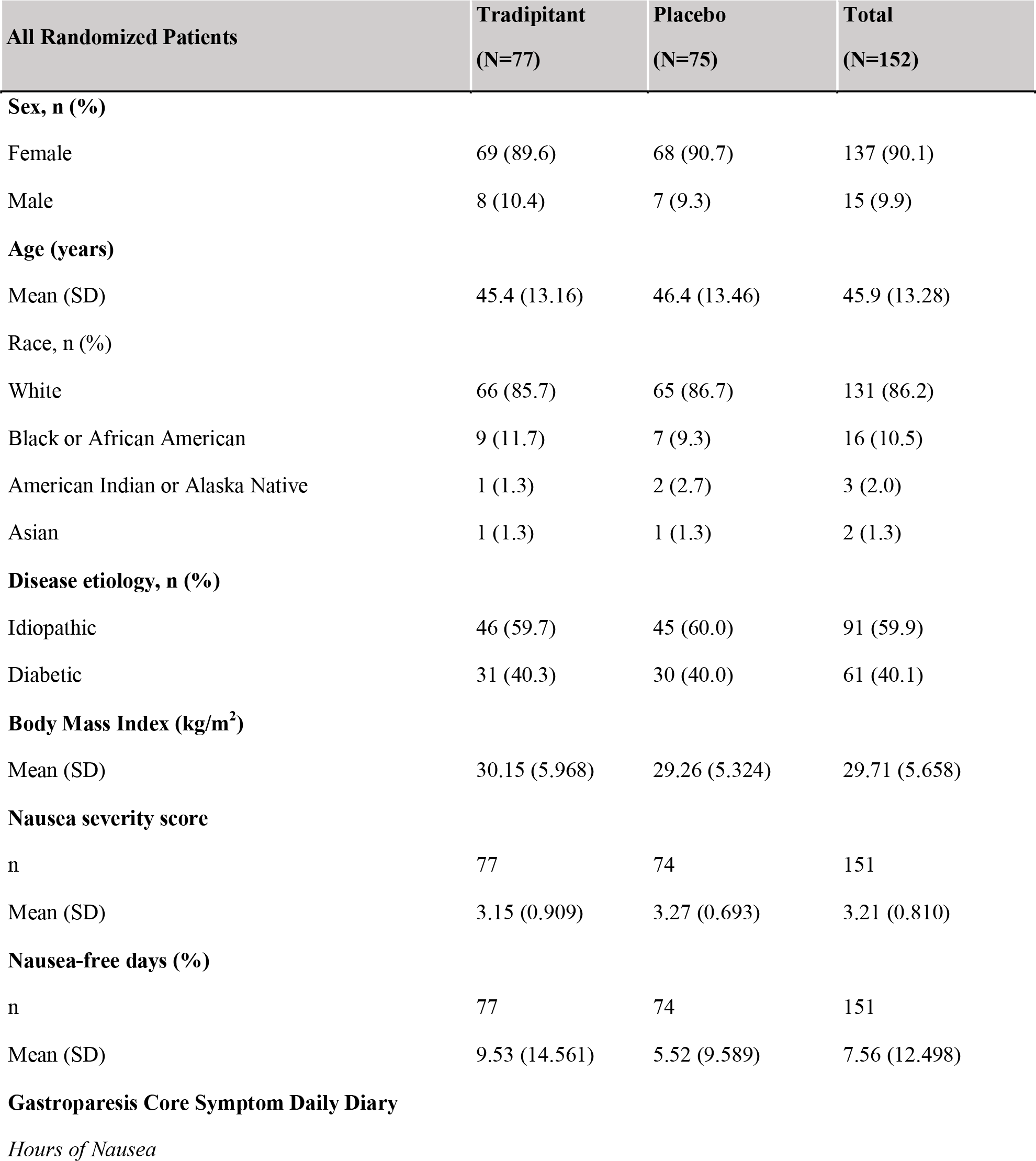
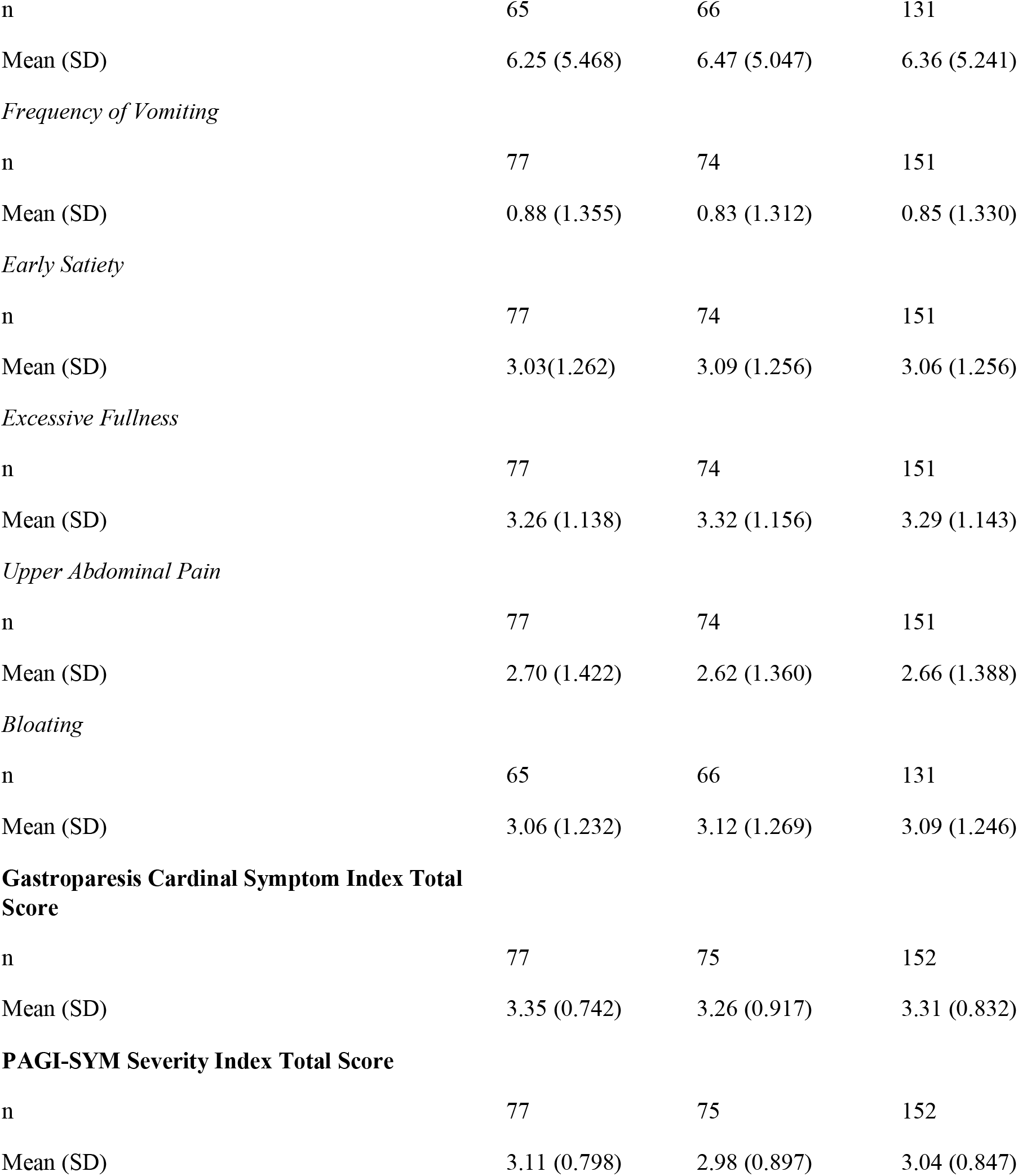
Key Study Demographics and Baseline Measures

| All Randomized Patients | Tradipitant<br>(N=77) | Placebo<br>(N=75) | Total<br>(N=152) |
| --- | --- | --- | --- |
| **Sex, n (%)** |  |  |  |
| Female | 69 (89.6) | 68 (90.7) | 137 (90.1) |
| Male | 8 (10.4) | 7 (9.3) | 15 (9.9) |
| **Age (years)** |  |  |  |
| Mean (SD) | 45.4 (13.16) | 46.4 (13.46) | 45.9 (13.28) |
| **Race, n (%)** |  |  |  |
| White | 66 (85.7) | 65 (86.7) | 131 (86.2) |
| Black or African American | 9 (11.7) | 7 (9.3) | 16 (10.5) |
| American Indian or Alaska Native | 1 (1.3) | 2 (2.7) | 3 (2.0) |
| Asian | 1 (1.3) | 1 (1.3) | 2 (1.3) |
| **Disease etiology, n (%)** |  |  |  |
| Idiopathic | 46 (59.7) | 45 (60.0) | 91 (59.9) |
| Diabetic | 31 (40.3) | 30 (40.0) | 61 (40.1) |
| **Body Mass Index (kg/m<sup>2</sup>)** |  |  |  |
| Mean (SD) | 30.15 (5.968) | 29.26 (5.324) | 29.71 (5.658) |
| **Nausea severity score** |  |  |  |
| n | 77 | 74 | 151 |
| Mean (SD) | 3.15 (0.909) | 3.27 (0.693) | 3.21 (0.810) |
| **Nausea-free days (%)** |  |  |  |
| n | 77 | 74 | 151 |
| Mean (SD) | 9.53 (14.561) | 5.52 (9.589) | 7.56 (12.498) |
| **Gastroparesis Core Symptom Daily Diary** |  |  |  |
| *Hours of Nausea* |  |  |  |
| n | 65 | 66 | 131 |
| Mean (SD) | 6.25 (5.468) | 6.47 (5.047) | 6.36 (5.241) |
| *Frequency of Vomiting* |  |  |  |
| n | 77 | 74 | 151 |
| Mean (SD) | 0.88 (1.355) | 0.83 (1.312) | 0.85 (1.330) |
| *Early Satiety* |  |  |  |
| n | 77 | 74 | 151 |
| Mean (SD) | 3.03(1.262) | 3.09 (1.256) | 3.06 (1.256) |
| *Excessive Fullness* |  |  |  |
| n | 77 | 74 | 151 |
| Mean (SD) | 3.26 (1.138) | 3.32 (1.156) | 3.29 (1.143) |
| *Upper Abdominal Pain* |  |  |  |
| n | 77 | 74 | 151 |
| Mean (SD) | 2.70 (1.422) | 2.62 (1.360) | 2.66 (1.388) |
| *Bloating* |  |  |  |
| n | 65 | 66 | 131 |
| Mean (SD) | 3.06 (1.232) | 3.12 (1.269) | 3.09 (1.246) |
| **Gastroparesis Cardinal Symptom Index Total Score** |  |  |  |
| n | 77 | 75 | 152 |
| Mean (SD) | 3.35 (0.742) | 3.26 (0.917) | 3.31 (0.832) |
| **PAGI-SYM Severity Index Total Score** |  |  |  |
| n | 77 | 75 | 152 |
| Mean (SD) | 3.11 (0.798) | 2.98 (0.897) | 3.04 (0.847) |

**Figure 1.**
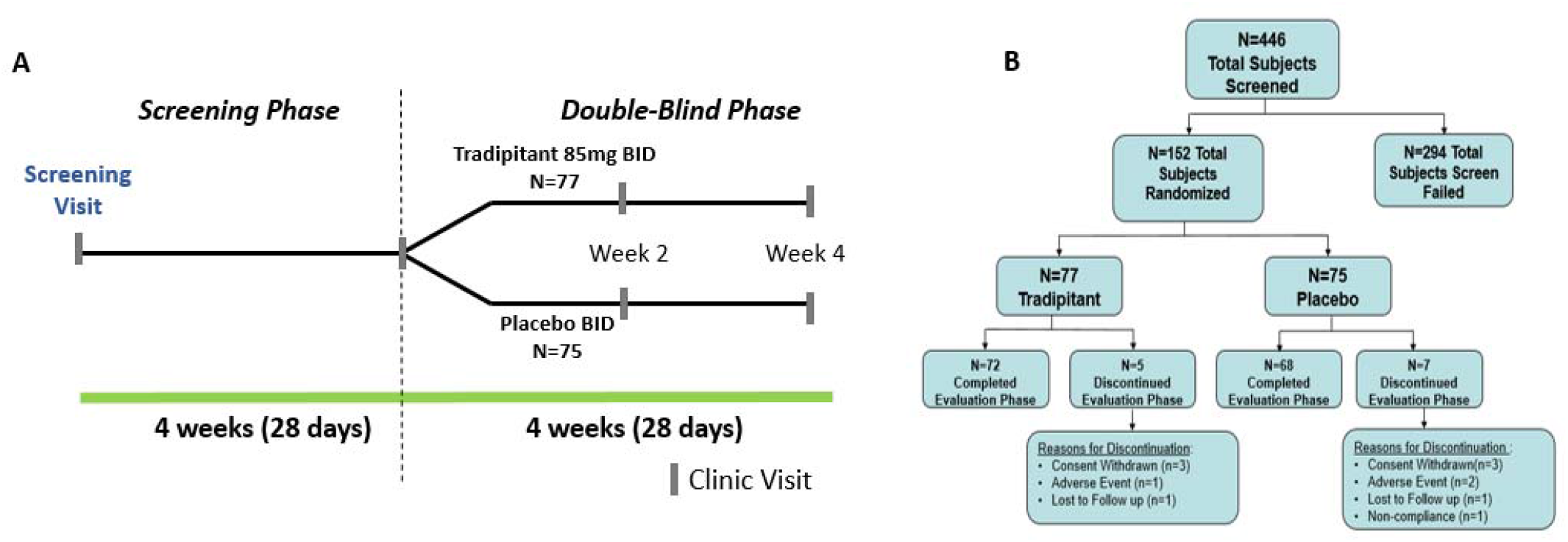
VP-VLY-686-2301 Study Design and Subject Flow Diagram.

The baseline nausea severity score was moderate to severe at 3.21 (± 0.810) and the baseline percentage of nausea free days was 7.56 % (± 12.498, Table 1). Baseline average scores of other core gastroparesis symptoms were moderate to severe in degree, however, every symptom was not present in every patient (Table 2).

**Table 2.** Baseline Measures in the Intention-to-Treat Population

| **ITT Population** | **Tradipitant** | **Placebo** |
| --- | --- | --- |
| **Gastroparesis Core Symptom** |  |  |
| **Nausea** |  |  |
| Mean (n) | 3.1 (73) | 3.2 (68) |
| Baseline >0 Mean (n)* | 3.1 (73) | 3.2 (68) |
| Percent of Symptom in Population (%) | 100% | 100% |
| **Vomiting** |  |  |
| Mean (n) | 0.9 (73) | 0.7 (68) |
| Baseline >0 Mean (n)* | 1.1 (58) | 1.2 (43) |
| Percent of Symptom in Population (%) | 79.5% | 63.2% |
| **Early Satiety** |  |  |
| Mean (n) | 3.0 (73) | 3.04 (68) |
| Baseline >0 Mean (n)* | 3.0 (73) | 3.1 (67) |
| Percent of Symptom in Population (%) | 100% | 98.5% |
| **Excessive Fullness** |  |  |
| Mean (n) | 3.2 (73) | 3.3 (68) |
| Baseline >0 Mean (n)* | 3.3 (72) | 3.3 (67) |
| Percent of Symptom in Population (%) | 98.6% | 98.5% |
| **Bloating** |  |  |
| Mean (n) | 3.1 (62) | 3.1 (61) |
| Baseline >0 Mean (n)* | 3.1 (62) | 3.2 (60) |
| Percent of Symptom in Population (%) | 100% | 98.4% |
| **Upper Abdominal Pain** |  |  |
| Mean (n) | 2.6 (73) | 2.6 (68) |
| Baseline >0 Mean (n)* | 2.7 (70) | 2.7 (66) |
| Percent of Symptom in Population (%) | 95.9% | 97.1% |

\*For patients with baseline score > 0

### Primary effects on nausea

The primary end point of this study was change from baseline in average nausea severity at Week 4 (0-5 scale). Tradipitant met the primary endpoint by significantly reducing nausea severity compared to patients receiving placebo (−1.25 vs. −0.73, p=.0099) (Table 3). Patients receiving tradipitant also reported a greater percentage of nausea-free days compared to patients receiving placebo (28.81% vs. 15.00%, p=0.0160). A numerical improvement in nausea severity was present by Week 2 and reached statistical significance by Week 3 through Week 4 (Figure 2A).

**Table 3.** Efficacy Measures in the Intention-to-Treat Population

**ITT Population (n=141)**
| Change from Baseline | Tradipitant n=73 | Placebo n=68 | P-value |
| --- | --- | --- | --- |
| **GCS-DD** |  |  |  |
| *Nausea Severity (at week 4)* | -1.25 | -0.73 | 0.0099 |
| *Nausea Free Days (%)* | 28.8 | 15.0 | 0.0160 |
| **GCSI** | -0.93 | -0.58 | 0.0223 |
| **PAGI-SYM** | -0.93 | -0.65 | 0.0497 |
| **CGI-Severity** | -1.13 | -0.74 | 0.0207 |
| **PGI-Change** | 2.66 | 3.06 | 0.0429 |

**Figure 2.**
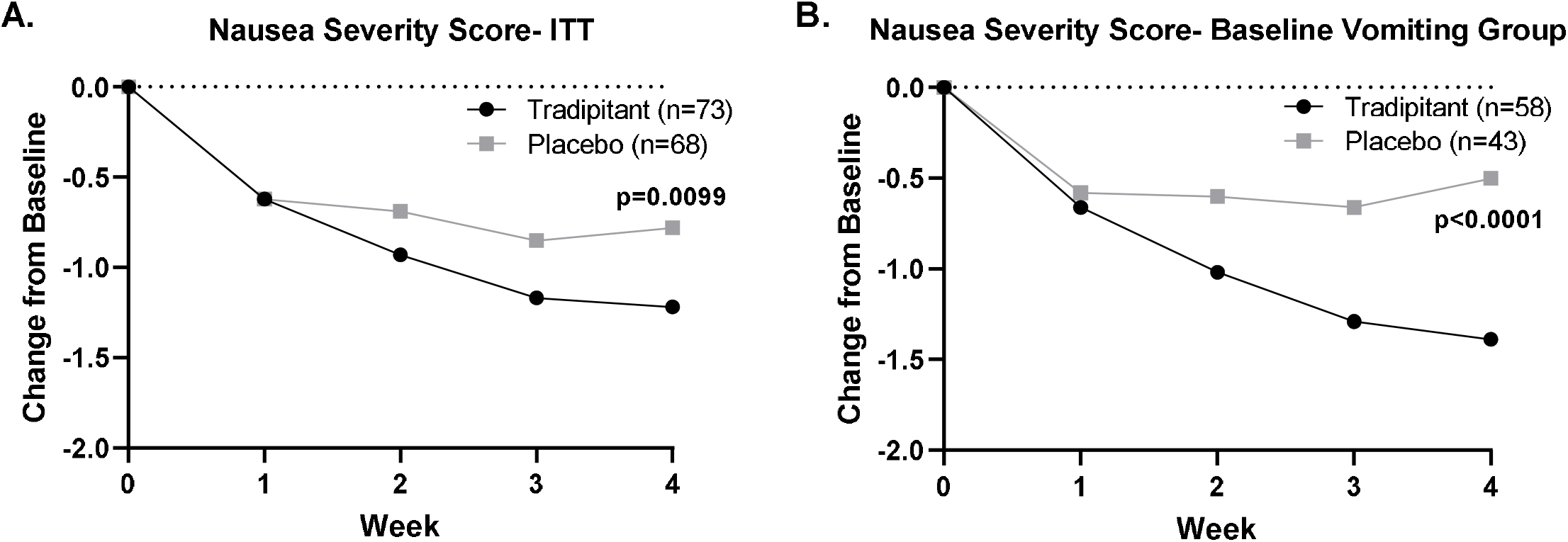
GCSDD Nausea Severity Score mean change-from-baseline for the placebo and tradipitant dose groups. Mean Values of changes from baseline during treatment with tradipitant and placebo for up to 4 weeks are shown along with p-values for overall treatment effect. A) Change over time of average nausea score in the ITT population during treatment with tradipitant (black, n=73) and placebo (gray, n=68). B) Change in time of the Baseline Vomiting Group population during treatment with tradipitant (black, n=58) and placebo (gray, n=43).

### Secondary and Post-hoc Endpoints

Tradipitant demonstrated significant improvement in most secondary endpoints studied, including key scales reflecting overall gastroparesis symptoms; GCSI (p=0.0223); PAGI-SYM (p=0.0497); CGI-S (p=0.0207); and PGI-C (p=0.0429) (Table 3). The change from baseline in GCSI total score to Week 4 (score at Day 28) was −0.93 for the tradipitant group compared to −0.58 for the placebo group and was statistically significant (p=0.022) (Table 3). Improvements were seen in most of the core gastroparesis symptoms. Tradipitant significantly improved average vomiting frequency and demonstrated a numerical improvement in the ability to finish a meal, excessive fullness, bloating, and upper abdominal pain compared placebo (Figure 3A). The change from baseline to Week 4 (Day 22 to 28) in daily average vomiting frequency was −0.49 for tradipitant vs. −0.26 for placebo and was statistically significant (p=0.039) (Figure 3A).

**Figure 3.**
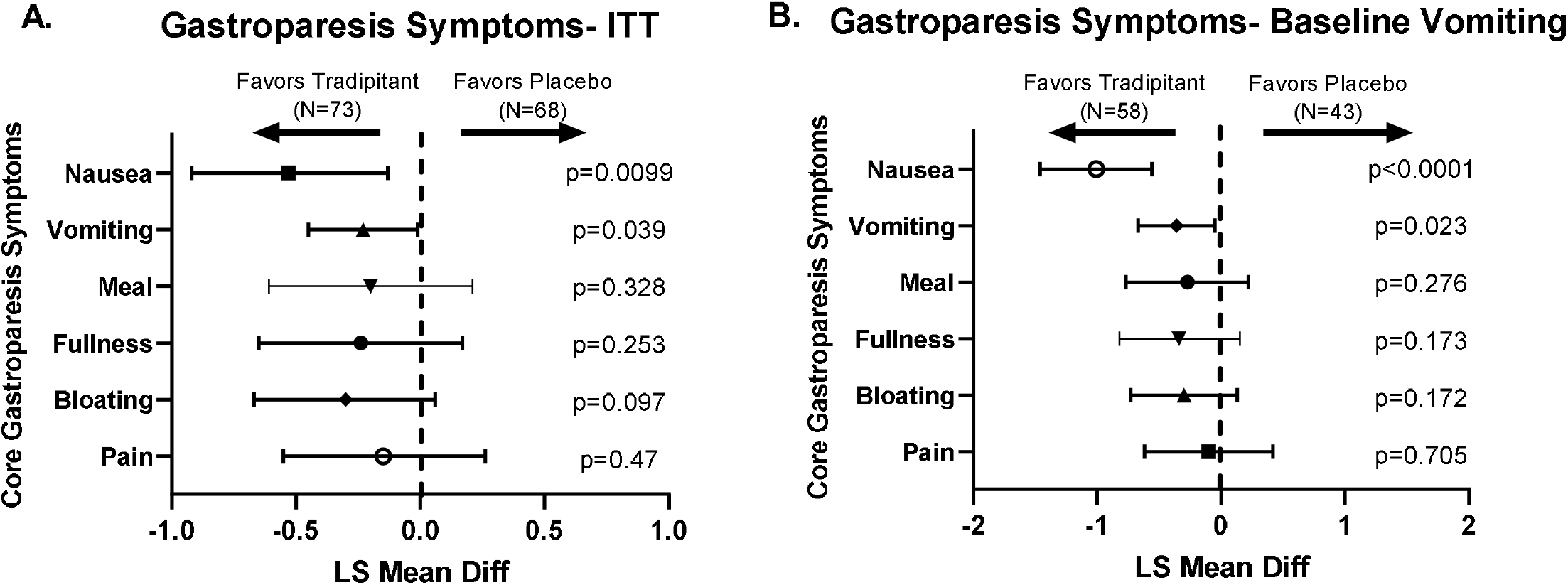
GCSDD Nausea Severity Score mean change-from-baseline for the placebo and tradipitant dose groups by disease etiology. Mean Values of changes from baseline during treatment with tradipitant and placebo for up to 4 weeks are shown along with p-values for overall treatment effect for both diabetic and idiopathic gastroparesis groups. A) Change over time of average nausea score in diabetic gastroparesis population during treatment with tradipitant (black, n=28) and placebo (gray, n=27). B) Change over time of average nausea score idiopathic gastroparesis population during treatment with tradipitant (black, n=45) and placebo (gray, n=41).

### Baseline Vomiting Subgroup

Patients in Baseline Vomiting Group (n=101) who received tradipitant had a significantly greater decrease in nausea severity compared to patients who received placebo (−1.43 vs −0.42, p<0.0001) (Table 4). Patients in this subgroup who received tradipitant reported significantly more nausea-free days compared to patients who received placebo (32.3% vs 7.6%, p=0.0003) (Table 4). Daily average vomiting frequency significantly decreased in the tradipitant group compared to placebo, consistent with the ITT Population. The change from baseline in vomiting frequency was −0.69 for tradipitant vs. −0.32 placebo (p=0.023) (Figure 3B). The change from baseline in GCSI total score in the Baseline Vomiting Group was also significantly greater in patients receiving tradipitant compared to patients receiving placebo (−1.10 vs. −0.60 p=0.0078) (Table 4).

**Table 4.** Efficacy Measures in the Baseline Vomiting Group

**Vomiting Population (n=101)**
|  | **Tradipitant n=58** | **Placebo n=43** | **P-value** |
| --- | --- | --- | --- |
| **GCSDD** |  |  |  |
| *Nausea* | -1.43 | -0.42 | <0.0001 |
| *DD-% Nausea Free Days* | 32.3 | 7.6 | 0.0003 |
| **GCSI** | -1.10 | -0.60 | 0.0078 |
| **PAGI-SYM** | -1.06 | -0.69 | 0.0294 |
| **CGI-Severity** | -1.24 | -0.79 | 0.0229 |
| **PGI-Change** | 2.52 | 3.24 | 0.0018 |

### Diabetic and Idiopathic Subgroups

Diabetic and idiopathic subgroups had similar response to treatment. In the diabetic group, change from baseline in nausea severity was −1.12 for patients on tradipitant compared to −0.80 for patients on placebo and did not reach statistical significance (p=0.308) (Figure 4A). In the idiopathic group, change from baseline in nausea score was −1.30 for tradipitant compared to − 0.69 for placebo and was statistically significant (p=0.025) (Figure 4B).

**Figure 4.**
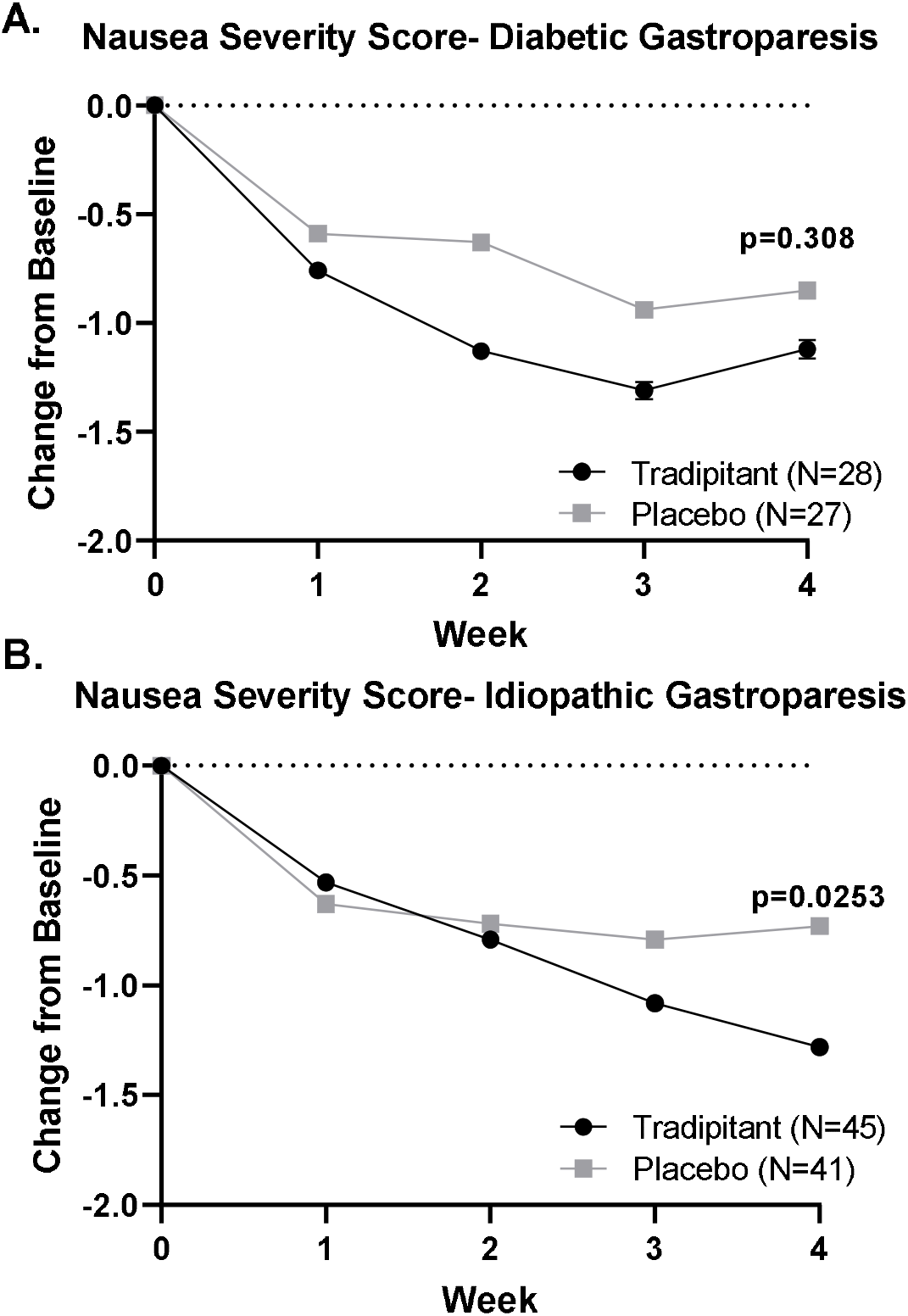
Forest Plot of the Analysis of Gastroparesis Core Symptom Daily Severity Scores in the ITT and Baseline Vomiting Group. A) Core Gastroparesis Symptoms of nausea, vomiting, ability to finish a meal (Meal), excessive fullness (Fullness), bloating, and upper abdominal pain (Pain) plotted as LS Mean Difference and 95% Confidence Interval after tradipitant or placebo treatment in the ITT Population. B) Core Gastroparesis Symptoms of nausea, vomiting, ability to finish a meal (Meal), excessive fullness (Fullness), bloating, and upper abdominal pain (Pain) plotted as LS Mean Difference and 95% Confidence Interval after tradipitant or placebo treatment in the Baseline Vomiting Population.

### Clinical Meaningfulness

A larger percentage of patients on tradipitant were considered nausea responders (defined as an average nausea score of “very mild” or better (≤ 1) at Week 4 days 22-28). 32.9% of patients on tradipitant compared to 11.8% of patients on placebo were nausea responders and this was statistically significant (p=0.0013, Figure 5A). 15.07% of patients treated with tradipitant were complete nausea responders (defined as having zero nausea at week 4) compared to 4.4% of patients on placebo (p=0.018, Figure 5C). 46.6% of patients treated with tradipitant had a clinically meaningful improvement on their total GCSI score (i.e., 1 point of greater improvement in their GCSI total score from baseline day 0 to week 4, day 28) compared to 23.5% of patients on placebo (p=0.0053 Figure 5E).

**Figure 5.**
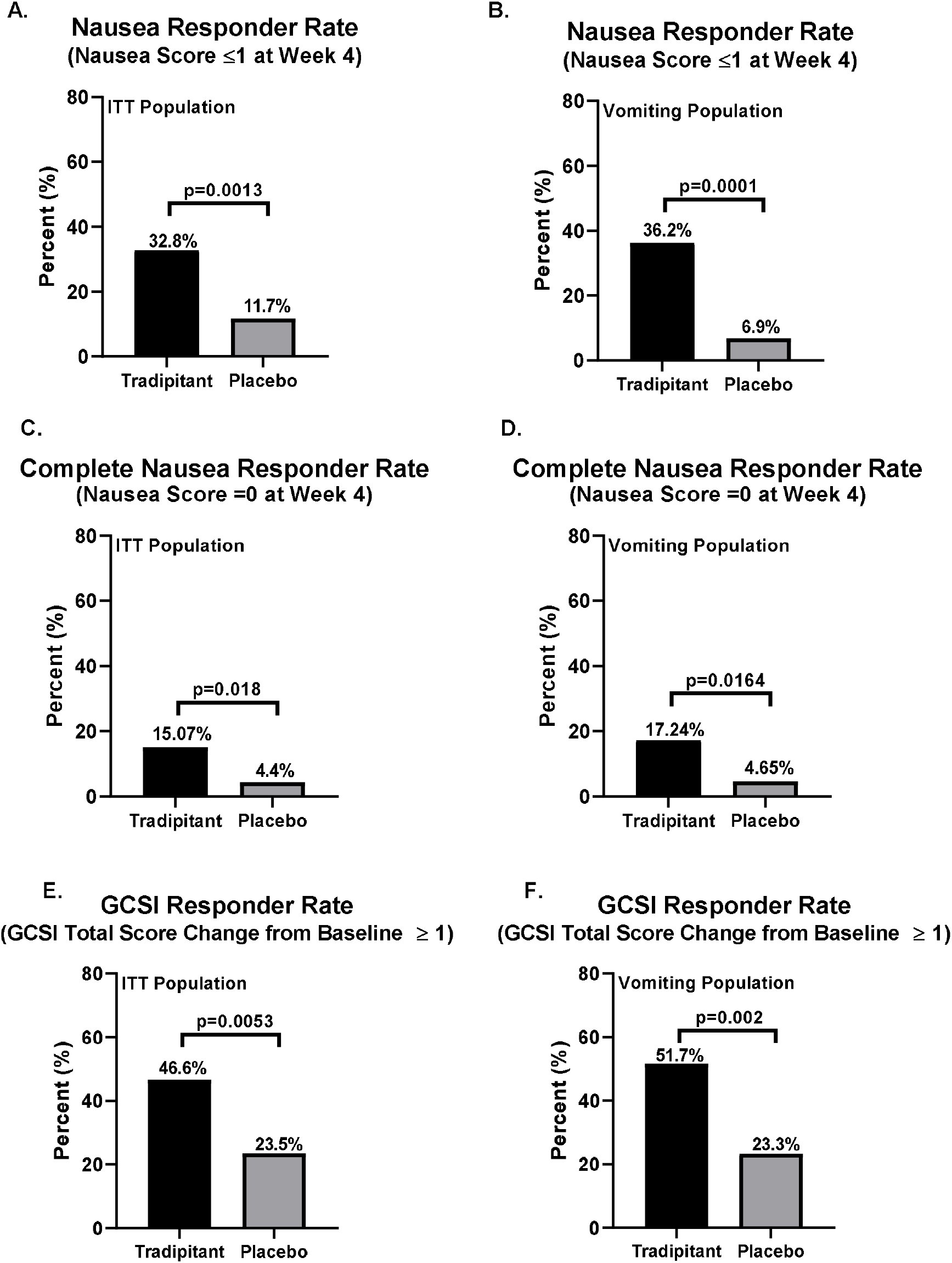
Clinically Meaningful responder analyses in ITT Population and Baseline Vomiting Group. A GCSI responder was defined as a subject having 1 or greater point improvement in GCSI total score at week 4. A) GCSI Total Score responder percentage in the ITT Population in tradipitant (black) and placebo (gray) groups. B) GCSI Total Score responder percentage in the Baseline Vomiting Population for tradipitant (black) and placebo (gray) groups. A complete nausea responder was defined as a subject averaging 1 (1= very mild) or less for daily nausea score over the last week of treatment. C) Nausea Responder percentage in the ITT Population in tradipitant (black) and placebo (gray) groups. D) Nausea Responder (Week 4 ≤ 1) percentage in the Baseline Vomiting Population for tradipitant (green) and placebo (blue) groups. E) Complete Nausea Responder (Week 4 = 0) percentage in the ITT Population in tradipitant (black) and placebo (gray) groups. F) Complete Nausea Responder percentage in the Baseline Vomiting Population for tradipitant (black) and placebo (gray) groups.

A larger percentage of patients on tradipitant in the Baseline Vomiting Group were also considered to have a meaningful response. 36.2% of patients treated with tradipitant were nausea responders in the Baseline Vomiting Group compared to 6.9% of patients on placebo (p=0.0001, Figure 5B). 17.24% of patients treated with tradipitant were complete nausea responders in the Baseline Vomiting Group compared to 4.65% of patients on placebo (p=0.0164, Figure 5D). 51.7 % of patients treated with tradipitant in this subgroup had a clinically meaningful improvement in their GCSI total score compared to 23.3% of patients on placebo (p=0.0020 Figure 5F).

### Minimal Important Difference (MID)

Using an anchor -based analysis, a 1 point PGI-C change from ‘No Change’ to ‘Minimal’ (a change from 4 to 3) in average nausea severity equaled a 0.506 improvement from baseline (Figure 6A). Based on these results, the average minimally important difference (MID) is 0.61 point change in nausea severity from baseline for a 1 point change on the PGI-C. For percentage of nausea free days, a 1 point change from ‘No Change’ to ‘Minimal’ (a change from 4 to 3) on the PGI-C is equal to a 7.8% improvement from baseline (Figure 6B). Based on these results, the average MID was calculated to be an improvement of 15.6% from baseline for a 1 point change on the PGI-C.

**Figure 6.**
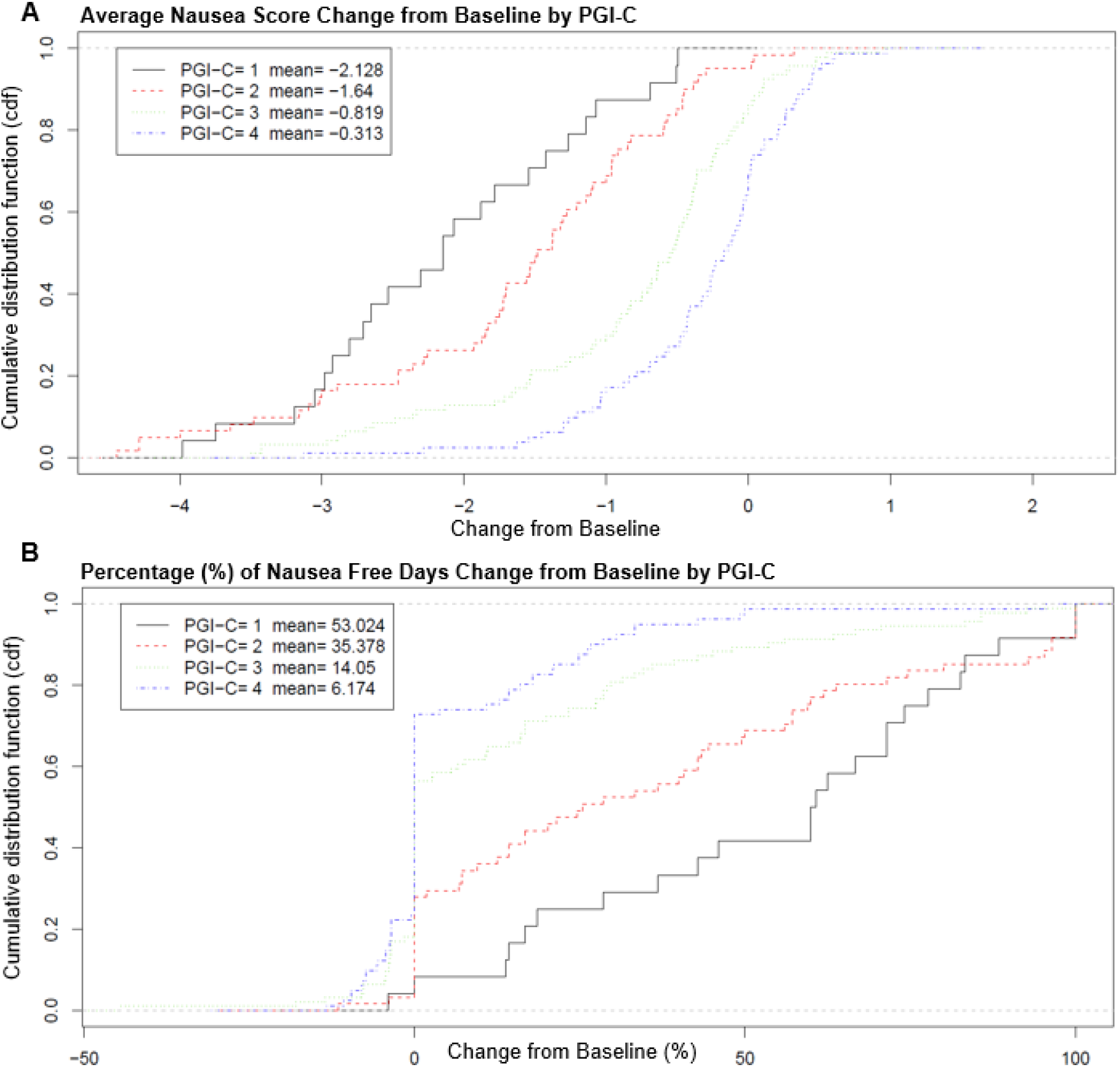
Anchor-based Analysis of Changes in PGI-C Score and Nausea Severity and Percentage of Nausea Free Days. Cumulative distribution function analysis of change from baseline in PGI-C and nausea severity. A) The correlation of 1-point changes in PGI-C scores with the change in nausea severity scores from baseline to Week 4. B) The correlation of 1-point changes PGI-C scores with the change in percentage of nausea free days from baseline to Week 4.

### Relationship between improvement in nausea and other symptoms

Patients who were nausea responders (nausea severity score ≤ 1 at Week 4) showed improvement on all symptoms present at baseline, and some of them reached statistical significance despite the small sample size (Figure 7A). Nausea responders had a LS mean difference of −2.12 in nausea severity score compared to placebo and was significant (p<0.0001, Figure 7A). Nausea responders also significantly improved in symptoms of early satiety (p=0.001), excessive fullness (p=0.001), bloating (p=0.013), and upper abdominal pain (p=0.0005) compared to placebo (Figure 7A).

**Figure 7.**
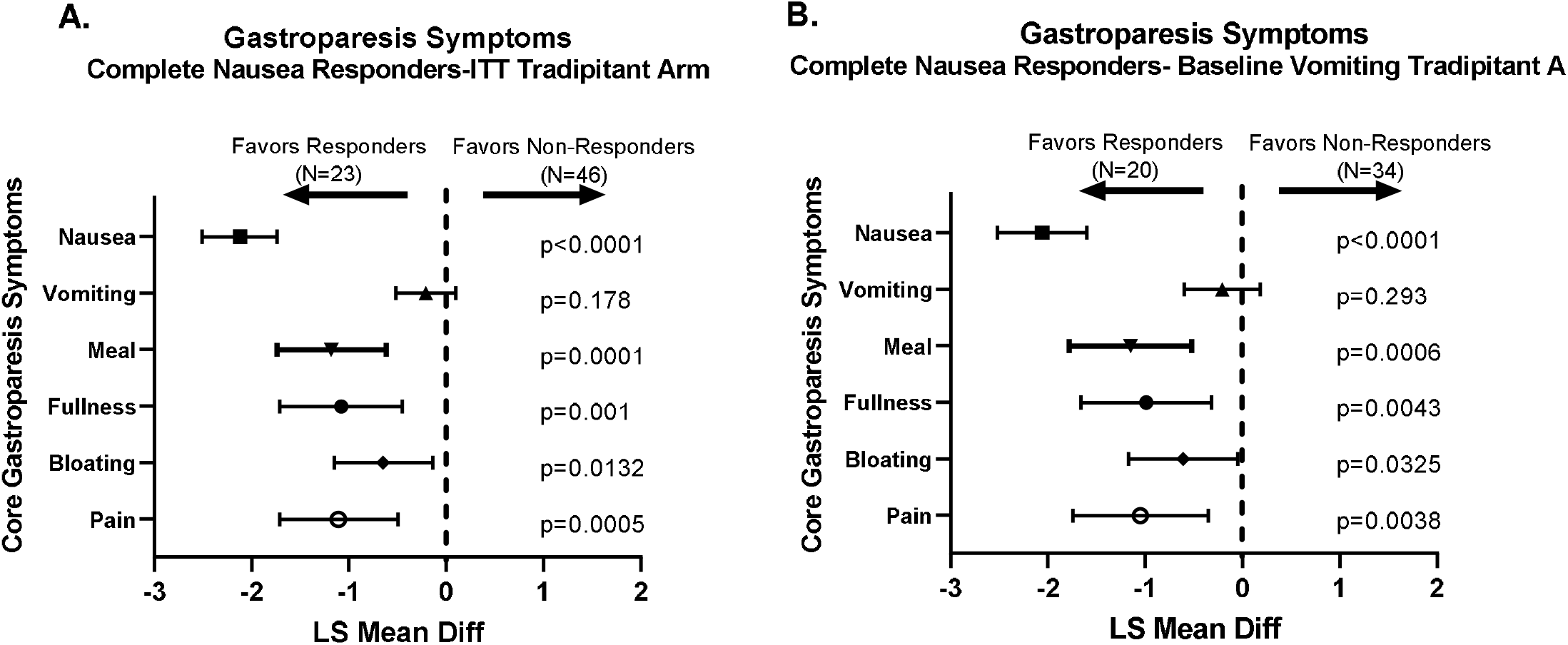
Forest Plot of the Analysis of Gastroparesis Core Symptom Daily Severity Scores by Nausea Responder. A) Core gastroparesis symptoms of nausea, vomiting, ability to finish a meal (Meal), excessive fullness (Fullness), bloating, and upper abdominal pain (Pain) plotted as LS Mean Difference and 95% Confidence Interval after treatment in the nausea responders and non-responders of the ITT Population. B) Analysis of the core gastroparesis symptoms of nausea, vomiting, ability to finish a meal (Meal), excessive fullness (Fullness), bloating, and upper abdominal pain (Pain) plotted as LS Mean Difference and 95% Confidence Interval after treatment in the nausea responders and non-responders of the Baseline Vomiting Group.

A similar analysis was performed in the Baseline Vomiting Group. Nausea responders in this subgroup on tradipitant had a LS mean difference of −2.06 in nausea severity score compared to placebo and this was significantly different (p<0.0001, Figure 7B). Nausea responders in the Baseline Vomiting Group also had significant improvement in early satiety (p=0.0006), excessive fullness (p=0.0043), bloating (p=0.0325), and upper abdominal pain scores (p=0.0038) in tradipitant treated patients compared to placebo (Figure 7B).

### Safety

Similar discontinuation rates and adverse event rates were seen between tradipitant and placebo groups. The most common were diarrhea, nausea, abdominal pain, dizziness, and headache. But these were rare and similar in tradipitant and placebo. The incidence of treatment emergent adverse events (TEAEs) was 31 (40.3%) in the tradipitant group compared to 20 (26.7%) in the placebo group (Table 5). No deaths were reported in the study. There was no clinically relevant or safety concerns identified in this study.

**Table 5.**
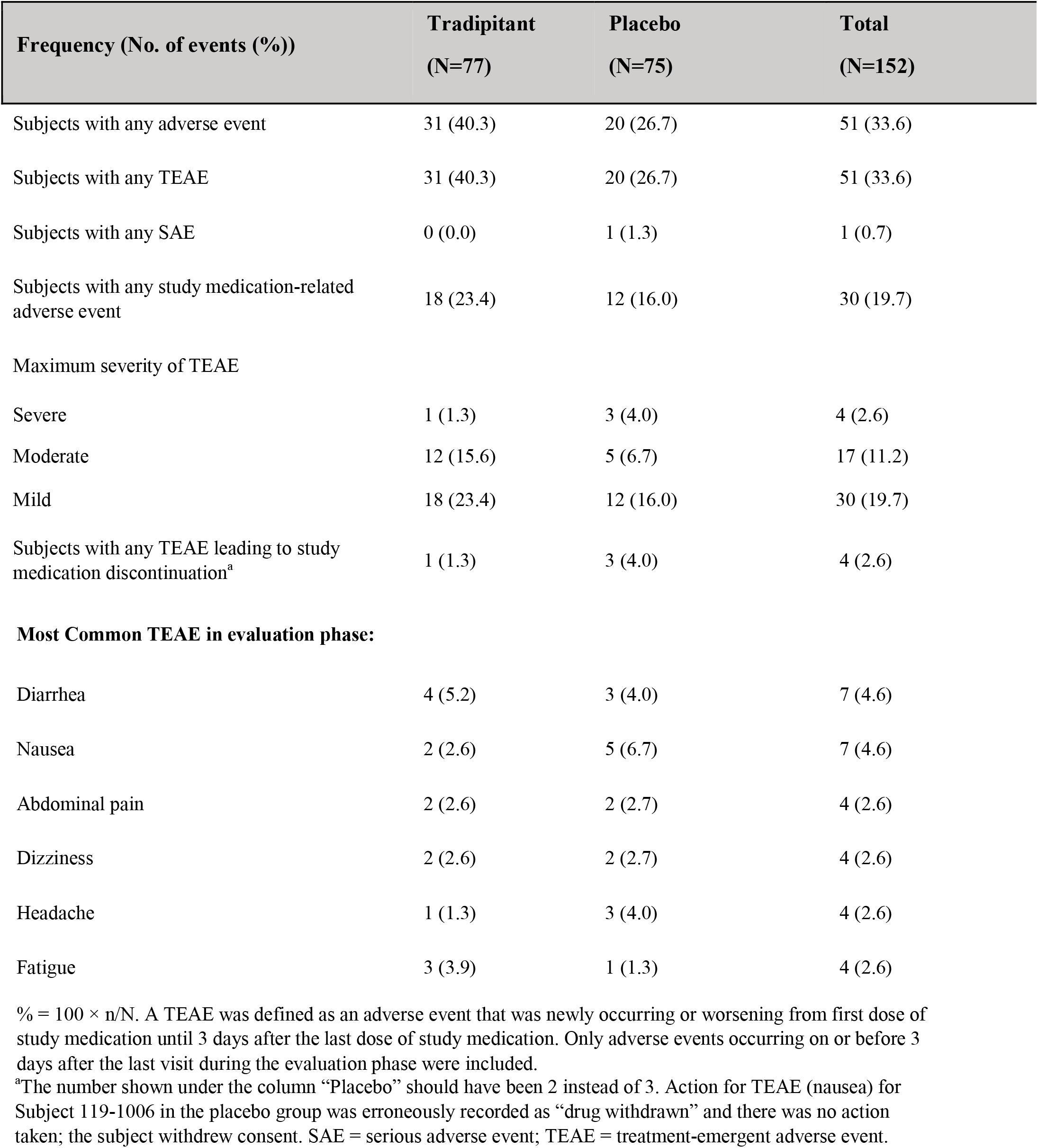
Frequency of reported Adverse Events by Treatment

| **Frequency (No. of events (%))** | **Tradipitant**<br>**(N=77)** | **Placebo**<br>**(N=75)** | **Total**<br>**(N=152)** |
| --- | --- | --- | --- |
| Subjects with any adverse event | 31 (40.3) | 20 (26.7) | 51 (33.6) |
| Subjects with any TEAE | 31 (40.3) | 20 (26.7) | 51 (33.6) |
| Subjects with any SAE | 0 (0.0) | 1 (1.3) | 1 (0.7) |
| Subjects with any study medication-related adverse event | 18 (23.4) | 12 (16.0) | 30 (19.7) |
| Maximum severity of TEAE |  |  |  |
| Severe | 1 (1.3) | 3 (4.0) | 4 (2.6) |
| Moderate | 12 (15.6) | 5 (6.7) | 17 (11.2) |
| Mild | 18 (23.4) | 12 (16.0) | 30 (19.7) |
| Subjects with any TEAE leading to study medication discontinuation<sup>a</sup> | 1 (1.3) | 3 (4.0) | 4 (2.6) |
| **Most Common TEAE in evaluation phase:** |  |  |  |
| Diarrhea | 4 (5.2) | 3 (4.0) | 7 (4.6) |
| Nausea | 2 (2.6) | 5 (6.7) | 7 (4.6) |
| Abdominal pain | 2 (2.6) | 2 (2.7) | 4 (2.6) |
| Dizziness | 2 (2.6) | 2 (2.7) | 4 (2.6) |
| Headache | 1 (1.3) | 3 (4.0) | 4 (2.6) |
| Fatigue | 3 (3.9) | 1 (1.3) | 4 (2.6) |

% = $100 \times n/N$ . A TEAE was defined as an adverse event that was newly occurring or worsening from first dose of study medication until 3 days after the last dose of study medication. Only adverse events occurring on or before 3 days after the last visit during the evaluation phase were included.
<sup>a</sup>The number shown under the column “Placebo” should have been 2 instead of 3. Action for TEAE (nausea) for Subject 119-1006 in the placebo group was erroneously recorded as “drug withdrawn” and there was no action taken; the subject withdrew consent. SAE = serious adverse event; TEAE = treatment-emergent adverse event.

## Discussion

In this 4-week Phase II randomized placebo-controlled trial involving patients with gastroparesis who reported moderate to severe nausea during screening period, tradipitant 85 mg BID demonstrated statistically significant and clinically meaningful improvement in nausea severity and overall gastroparesis compared to placebo. Tradipitant also significantly improved overall gastroparesis symptoms including frequency of vomiting, GCSI total score, PAGI-SYM total score, PGI-C and CGI-S. Tradipitant was effective in patients with baseline vomiting, and was equally effective in patients with diabetic and idiopathic gastroparesis. Importantly, tradipitant was well-tolerated with an adverse event profile similar to placebo.

A greater percentage of patients treated with tradipitant had a clinically significant improvement in nausea. Previous studies have calculated the minimally improved difference (MID) for nausea to be −0.55 as measured on the GCSI-DD^20^. The average nausea severity score improvement in this study was −1.25 for patients on tradipitant compared to −0.73 for patients on placebo (p=0.0099).

The improvement in percentage of nausea free days in the ITT Population was 28.8% for patients on tradipitant compared to 15.0% for patients on placebo (p=0.016). Comparing against the average MID calculated for nausea severity (0.61) and average MID calculated for nausea-free days (15.61%) in this study, the effect of tradipitant from baseline was approximately double these two calculated MIDs.

In this study, a greater percentage of patients had a clinically meaningful improvement in gastroparesis symptoms. Previous studies have reported a decrease in GCSI total score of 0.75^17^ and 1.0^18,19^ to be a clinically meaningful response. In this study, 46.6% and 23.5% of patients treated with tradipitant and placebo respectively had a clinically meaningful response to treatment as assessed by GCSI total score.

This magnitude of effect for tradipitant was large and clinically meaningful considering gastroparesis symptoms are persistent despite available treatments. A study of the natural history of diabetic gastroparesis showed upper GI symptoms in patients with diabetes were unchanging over 12 years with treatment^21^. Thus far, clinical trials comparing treatment to placebo failed to show as great of a percent change from baseline in daily average nausea, including aprepitant (−0.8 change after 4 weeks on a 5 point scale)^9^, and relamorelin (∼ −2.2 change after 12 weeks on a 10 point scale)^22^.

Subgroup analysis of the Baseline Vomiting Group showed a magnitude of improvement in nausea and overall gastroparesis symptoms was both statistically significant and clinically meaningful. This subgroup of patients (101/141) demonstrated numerically higher improvements in nausea severity scores, however this looks to be mainly driven by a smaller placebo effect. Additionally, this sub group demonstrated significant and clinically meaningful effects in change in the number of nausea-free days.

Tradipitant had similar efficacy in both diabetic gastroparesis and idiopathic gastroparesis. When analyzed as separate groups, the treatment effect in patients with idiopathic gastroparesis reached significance. The numeric improvement over placebo was slightly smaller in the diabetic group and did not reach significance, as the sample size was approximately half that of the idiopathic group and therefore lacked statistical power. Research on the symptomology of gastroparesis suggests nausea and vomiting may be more severe in diabetic gastroparesis than in idiopathic gastroparesis^23,24^; however; we did not observe that difference in our study, perhaps because only patients with moderate to severe nausea were enrolled as an inclusion criteria in this study.

The improvement in nausea seen in this trial is particularly notable because nausea is the most common symptom of gastroparesis (reported in more than 90% of patients) and is the most disruptive. Increasing severity of nausea is associated with impaired quality of life as measured on the PAGI-QOL.^24^ Patients in the current trial had moderate to severe nausea (i.e., a mean nausea score during the screening period of ≥ 3 on a 5=point scale). Despite the severity of baseline nausea, the average improvement in nausea was 1.25 compared with 0.73 in patients receiving placebo. In addition, patients receiving tradipitant had more days that were nausea free and had a higher percentage that were nausea responders (i.e., decrease of > 1). The number needed to treat (NNT) to achieve a nausea response of mild or zero nausea by week 4 was calculated to be approximately 3 to 4.

It was not an inclusion criteria for patients to present with the other core gastroparesis symptoms, however; over 95% of patients presented with additional core gastroparesis symptoms (having an average > 0 during screening). Numerical improvements were seen in most of the core gastroparesis symptoms as measured by the daily diary, including nausea, vomiting, bloating, and fullness after meals, consistent with an overall improvement and no associated worsening of any of the core symptoms.

The cause of nausea in gastroparesis is not well understood. Several neurotransmitter pathways are thought to be involved in activation of nausea in gastroparesis, including serotonin 5-HT3 and NK-1 mechanisms. Serotonin 5-HT3, Muscarinic M1, Histaminergic H1, and dopamine D2 receptor (D2) systems are already being studied for treatment of gastroparesis and other indications that involve nausea^25^. NK1Rs are expressed on enteric neurons, interstitial cells of Cajal, epithelial cells, and on the lymphocytes and macrophages of the lamina propria. NK1Rs are thought to regulate neuro-transmission, motility, secretion, inflammation, and pain in the gastrointestinal tract. NK-1R as a target for gastroparesis was studied in clinical trials by the Gastroparesis Clinical Research Consortium (Gp-CRC) with the NK-1R antagonist aprepitant^9^. This novel mechanism for treatment of gastroparesis failed to reach the primary endpoint of improvement in nausea score as measured by Visual Analog Scale (VAS). However, aprepitant did show improvement in nausea and other GI symptoms as measured by the GCSI. Aprepitant has also been found effective in a case of severe vomiting in a patient with diabetes^26^ and a case of long term refractory diabetic gastroparesis^27^. The results of this phase 2 study further validate the NK-1 receptor as a useful target for symptom relief in diabetic and idiopathic gastroparesis.

Improvement of nausea could possibly be mediated by acceleration in gastric emptying; however, gastric emptying was not measured as an endpoint in this trial. Blocking NK-1R in smooth muscle could increase gastric emptying, as it has been reported that aprepitant can increase gastric accommodation volume and maximum calorie volume tolerated in healthy volunteers^28^. Delayed emptying is a predictor of more severe upper gastrointestinal symptoms^29,6^; however, improvement in gastric emptying rate does not always correlate well with improvement in symptoms^22,30,31,32,33^. The effect of gastric emptying will be assessed in future studies, although we predict tradipitant to be useful for treatment of gastroparesis with or without an effect on gastric emptying rate.

Other remaining questions include not only the effect on gastric emptying, and the effect on gastric accommodation, but also the maintenance effect of long term tradipitant use. Gastroparesis is a chronic disease and efficacy of treatment would have to be safe and effective beyond 4 weeks to be useful to this population. There is some evidence of agonist induced receptor internalization of NK-1R^34^ and inflammation induced internalization of NK-1R^35^; however, we would not expect receptor internalization after tradipitant and therefore do not expect tachyphylaxis of effect in long term studies. Studies in tradipitant over 4 weeks are ongoing.

In conclusion, tradipitant met the primary pre-specified aim of this study and achieved clinically meaningful outcomes in patients with gastroparesis. Tradipitant was demonstrated to be well-tolerated with an adverse event profile similar to placebo. The overall benefit risk profile, if confirmed, is likely to offer advantages over both approved and off label treatments currently utilized. The effect of tradipitant in achieving complete response in nausea but also improving overall symptoms may suggest a disease modifying effect through an action to the local neuromuscular network as well as the central nervous system centers for nausea and vomiting.

## Data Availability

Authors had access to the study data and had reviewed and approved the final manuscript.

## Acknowledgements

We would like to acknowledge the contributions of Dr. Pankaj Jay Pasricha who worked as a consultant on this project. We would also like to acknowledge the Investigators and the 47 contributing centers that participated in conducting this study.

## Abbreviations

list abbreviations (in alphabetical order) not mentioned in the Style Guide following the Instructions to Authors. (Note: In general, the use of abbreviations is discouraged).

ALT: alanine aminotransferase
ANMS GCSI-DD: American Neurogastroenterology Motility Society Gastroparesis Cardinal Symptom Index Daily Diary
AST: aspartate aminotransferase
BID: Bis in die (Latin two times a day dosing)
CGI-S: Clinician Global Impression of Severity
GCSI: Gastroparesis Cardinal Symptom Index
GCSDD: Gastroparesis Core Symptom Daily Diary
ITT: Intent-to-Treat
LS Mean: Least Squared Mean
NK-1R: Neurokinin-1 Receptor
NNT: Number needed to treat
PAGI-SYM: Patient Assessment of Gastrointestinal Disorders Symptom Severity Index
PAGI-QOL: Patient Assessment of Gastrointestinal Disorders-Quality of Life
PGI-C: Patient Global Impression of Change
SP: Substance P

